# Clinically distinct COVID-19 cases share strikingly similar immune response progression: A follow-up analysis

**DOI:** 10.1101/2020.09.16.20115972

**Authors:** Melissa A. Hausburg, Kaysie L. Banton, Michael Roshon, David Bar-Or

## Abstract

Inflammatory responses to the novel coronavirus SARS-CoV-2, which causes COVID-19, range from asymptomatic to severe. Here we present a follow-up analysis of a longitudinal study characterizing COVID-19 immune responses from a father and son with distinctly different clinical courses. The father required a lengthy hospital stay for severe symptoms, whereas his son had mild symptoms and no fever yet tested positive for SARS-CoV-2 for 29 days. Father and son, as well as another unrelated COVID-19 patient, displayed a robust increase of SERPING1, the transcript encoding C1 esterase inhibitor (C1-INH). We further bolstered this finding by incorporating a serum proteomics dataset and found that serum C1-INH was consistently increased in COVID-19 patients. C1-INH is a central regulator of the contact and complement systems, potentially linking COVID-19 to complement hyperactivation, fibrin clot formation, and immune depression. Furthermore, despite distinct clinical cases, significant parallels were observed in transcripts involved interferon and B cell signaling. As symptoms were resolving, widespread decreases were seen in immune-related transcripts to levels below those of healthy controls. Our study provides insight into the immune responses of likely millions of people with extremely mild symptoms who may not be aware of their infection with SARS-CoV-2 and implies a potential for long-lasting consequences that could contribute to reinfection risk.

## Introduction

Coronavirus disease 2019 (COVID-19), which is caused by infection with the novel coronavirus SARS-Coronavirus-2 (SARS-CoV-2), was first described in Wuhan, China, at the end of 2019. COVID-19 and SARS-CoV-2 infections rapidly spread, and COVID-19 was declared a pandemic by the World Health Organization in March of 2020.

The response to the COVID-19 pandemic by the scientific community has resulted in a concentrated effort of historical proportions, including widespread data sharing in the hopes of understanding every aspect of SARS-CoV-2 and COVID-19. Symptoms of SARS-CoV-2 infection are varied, as some patients are asymptomatic, whereas others suffer from cough, dyspnea, respiratory failure, cytokine storm, and, in many cases, death (1). Furthermore, our understanding of COVID-19 symptoms continues to evolve as unexpected consequences of SARS-CoV-2 infection come to light, e.g., potential increased risk of thrombotic events in COVID-19 patients (2-6). SARS-CoV-2 infections spread through contact with symptomatic, as well as pre-symptomatic and asymptomatic, cases (7). A detailed characterization of immune responses to SARS-CoV-2 infection in pre-symptomatic, mild, and, asymptomatic cases is warranted and imperative for the identification of predictive biomarkers and effective vaccination programs.

Our goal was to apply statistical comparisons to the three clinical case subjects from Wuhan, China, presented by Ong et al., as the original paper was not subjected to statistical analysis (8). These data are unique because cases 1 and 2 are father and son with starkly different clinical courses of COVID-19 infection. These two cases represent a dataset with reduced genetic diversity and offer the ability to compare a severe to a mild case temporally, as Ong et al. characterized whole blood mRNA ranging from early in the clinical course to as late as 19 days from symptom onset (DSO)(8).

Case 1, a 66-year-old male, presented with fever (38.1 oC), cough, and, as defined by Ong et al., “bilateral, patchy, ill-defined lung infiltrates”(8). For the purposes of our study, we classified case 1 as a severe COVID-19 case, consistent with previous studies (9). At 5 DSO, when his arterial O2 saturation (SO2) reached a nadir, case 1 received the anti-viral medication lopinavir-ritonavir; however, anti-viral administration did not prevent case 1 from testing positive for SARS-CoV-2 at 7, 12, and 18 DSO. Case 1 was deemed fit for discharge on 22 DSO.

The son of case 1 was case 2, a 37-year-old who began to have COVID-19 symptoms just prior to the onset of symptoms in case 1. Case 2 reported having a single day of diarrhea and, two days later, a cough and mild sore throat. Ong et al. deemed day 1 of his clinical course as the day that the sore throat and cough symptoms began (8). The concept of DSO is subjective and, in cases 1 and 2, symptom onset was so closely reported that it is likely that father and son became inoculated with SARS-CoV-2 at, or closely around, the same time. The son self-reported a mild sore throat and cough on January 19, and the father self-reported a fever on January 20 (8). A fever is a quantitative measurement and indicates a systemic inflammatory response, and this was set as 1 DSO, in the case of the father. Complicating the situation, the son never reported or developed a fever. Assignment of 1 DSO was set at the time the son developed a mild sore throat and cough, one day prior to the father’s reported fever. The overall timing and relationship of father and son suggests these samples were closely matched in time of infection, regardless of the DSO reported in Ong et al. (8). Case 2’s cough persisted until 19 DSO, yet he tested positive for SARS-CoV-2 during 1-21 DSO and again on 23, 26, and 29 DSO, the latter being a full 10 days after symptom resolution. For these analyses, case 2 was deemed to be a mild case, as he was afebrile and never required medical support.

Case 3 was an unrelated 37-year-old man that presented to the hospital with fever, non-productive cough, lethargy, and myalgia and was confirmed positive for SARS-CoV-2 on 7 DSO. Throat swabs from case 3 tested positive for SARS-CoV-2 consistently until 17 DSO and intermittently until 23 DSO. We deemed his case to be moderate as he did not require any supplemental oxygen.

In a related study, Hadjadi et al. characterized whole blood mRNA from 32 COVID-19 patients separated into groups by disease severity, mild/moderate, severe, and critical (10), and herein we provide comparisons to these data. We also compare our findings to those of Shen et al. that performed a full proteomics and metabolomic analysis of COVID-19 patient serum (11).

By applying unbiased sample grouping and statistical methods, this dataset may provide a window into the temporal dynamics of COVID-19-related immune responses in afebrile patients with mild symptoms.

## Methods

### Hierarchical Clustering and PCA analysis

Data were generated by Ong et al. using the Human Immunology V2 multiplex panel available from NanoString Technologies, Inc (8). Raw data were obtained from Array Express, accession number E-MTAB-8871, and uploaded into the nSolver Analysis software (NanoString Technologies, Inc.) for further analysis. All data, analysis software, and patient information used in this study are freely available to the public. These data were not generated from patients at any of our facilities, and thus, institutional review board oversight was not necessary. Only case 1 day 4 was run in technical duplicates, and as no other sample was run in replicate, we removed the second replicate for this sample from any further analysis. Data were subjected to the Advanced Analysis Module of the nSolver software and were normalized using default settings (Figure 1). For heatmap generation, each line represents a single gene, each column is a separate sample, and means were scaled (z-score) to give all genes equal variance. The clustering dendrogram was generated unsupervised using Euclidean distance and complete linkage. Orange corresponds to higher expression, and blue corresponds to lower expression. Principal component analysis (PCA) plots were auto generated by nSolver Advanced Analysis software, NanoString, Technologies Inc.

**Figure 1.**
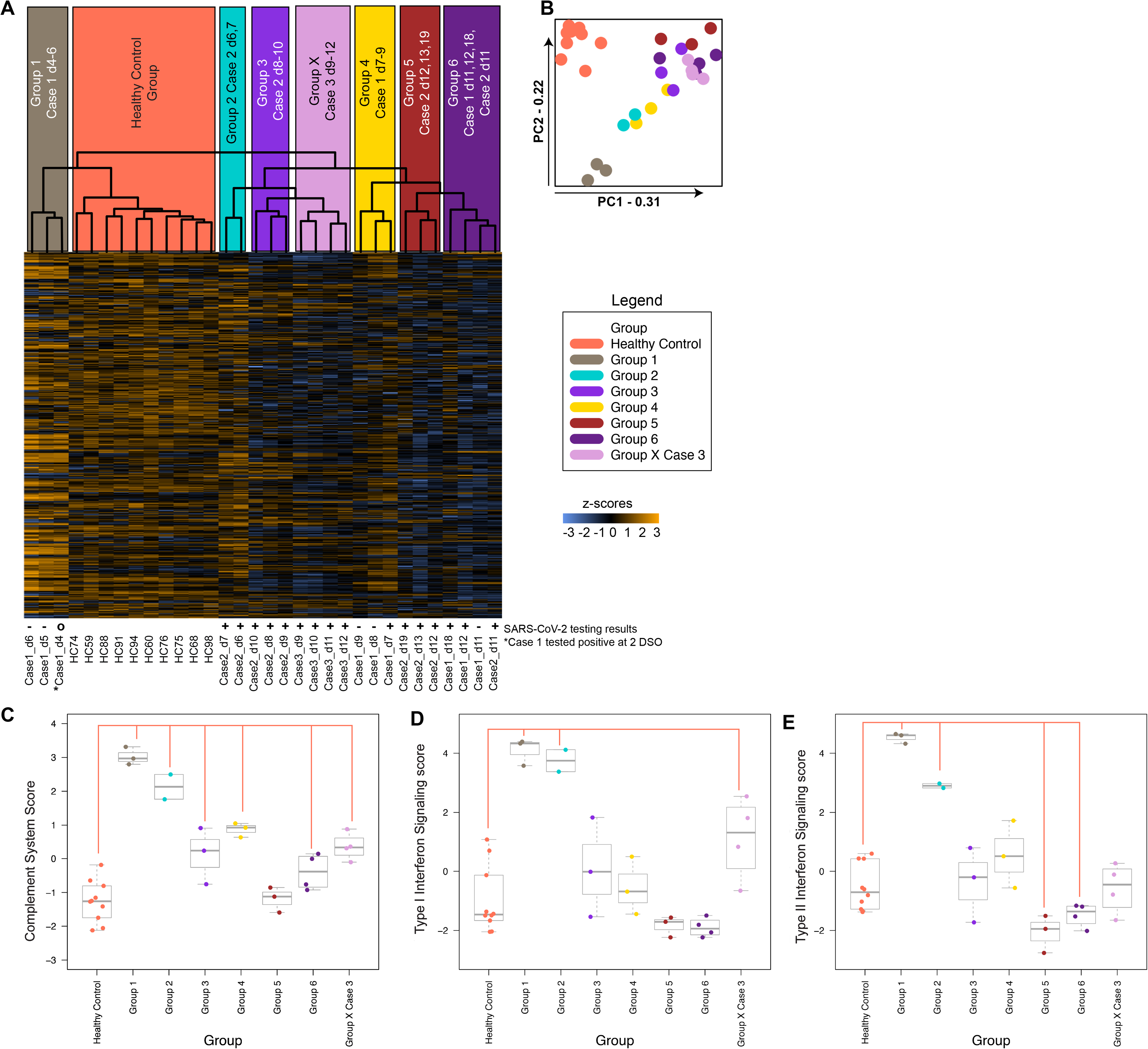
COVID-19 increased complement scores and elicited interferon type-I signaling response regardless of symptom severity. (A) Hierarchical clustering analysis dendrogram showing eight clusters over 32 samples that were used to form groups for further analysis. Orange indicates expression higher than the mean, and blue indicates expression lower than the mean. (B) Principal component (PC) analysis showing that groups defined based on unsupervised hierarchical clustering also associated with each other over PC1 and PC2. PC1 and PC2 contributed to 31% and 22%, respectively, of the variability in these data. (C-E) Boxplot graphs of samples showing the mean and variance of pathway scores plotted against each group defined in (A); salmon-colored bars indicate scores that are significantly different from HC (p<0.05). (C) Complement system scores showing significantly more positive scores compared to HC in groups 1-4, 6, and X. (D) IFN-I signaling pathway scores were significantly more positive in groups 1, 2, and X compared to HC. (E) Type II pathway scores were significantly more positive in groups 1, 2, 5, 6, and X compared to HC.

### Pathway Analysis

nSolver software condensed each covariate’s gene expression profiles into pathway scores. Pathway scores were fit using the first principal component of the data for each gene set (Figure 1). A positive score indicates that at least half of the genes in that pathway’s set have positive weights, and vice versa for negative scores. Pathway scores were subjected to gaussian statistical analysis t-tests with homogeneous variances.

### Differential Gene Expression Analysis

Differential gene expression analysis was performed with nSolver software using “groups” as the covariate (Figure 2). The Benjamini-Yekutieli method of FDR calculations was selected, and all other analyses were performed with the default settings.

**Figure 2.**
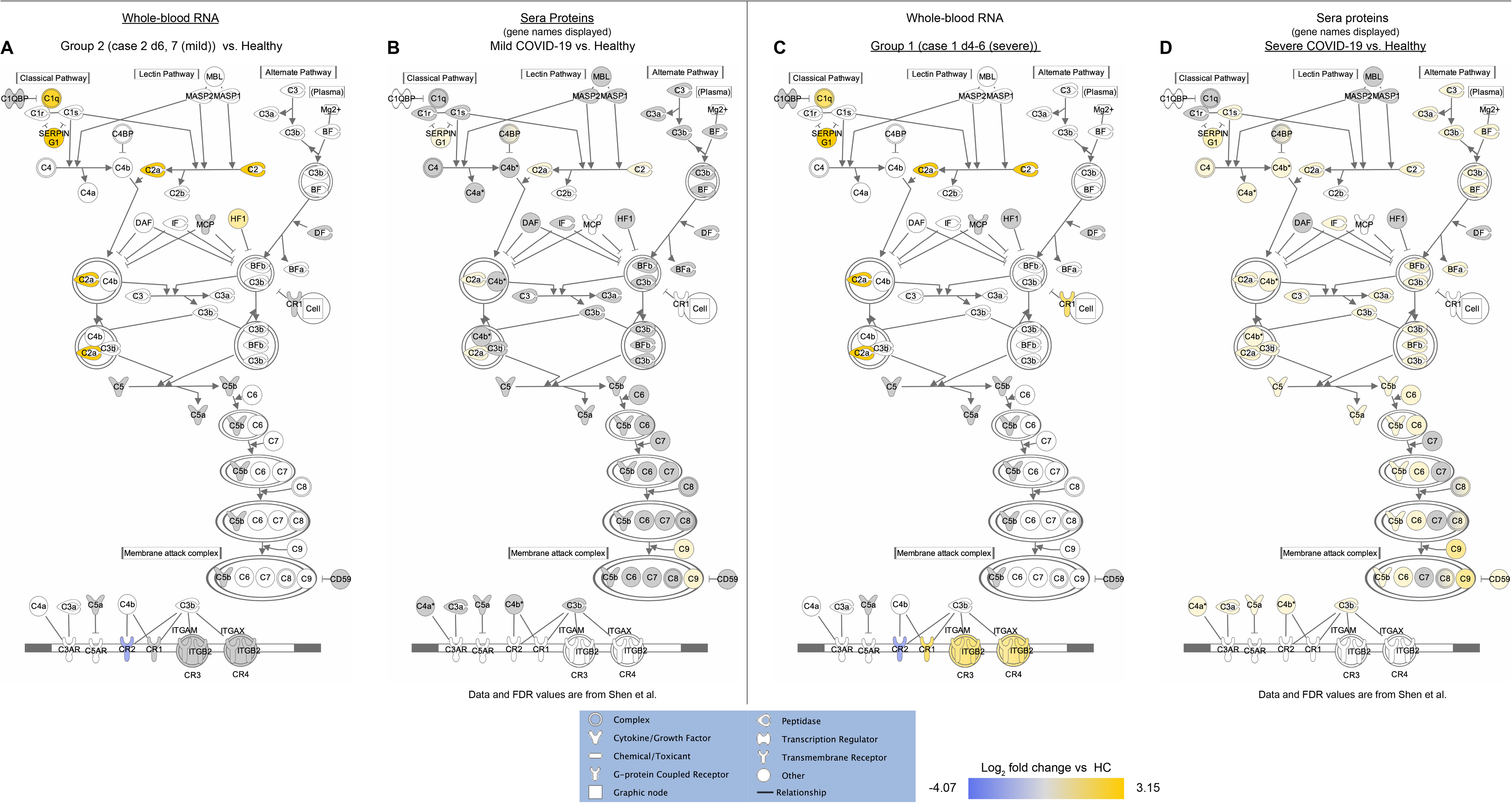
COVID-19 increased SERPING1 and the encoded protein, C1 esterase inhibitor, a central regulator of coagulation and complement regulation, regardless of symptom severity. (A-D) Ingenuity Pathway Analysis (IPA) “Complement System” canonical pathway; yellow and blue fill denotes significantly (FDR<0.01) increased or decreased log2 fold-regulation over HC, respectively. (A and C) Overlaid differential expression (DE) values of RNA when compared to HC for groups 2 and 1, respectively. Grayed molecules are in the dataset but do not meet the cutoff of more than 2-fold regulated with FDR<0.01. (B and D) Overlaid serum protein values for patients with non-severe and severe COVID-19, respectively, compared to healthy controls. Values and FDR calculations were performed as in Shen et al. (11). The corresponding gene names are displayed, although these data represent differentially abundant serum proteins. Grayed molecules are in the dataset but do not meet the cutoff of FDR<0.01. The key to each gene name can be found in Supplemental Table 5.

To compare our findings to the related study by Hadjadi et al. we obtained the normalized, log-transformed values from the authors (10). For differential gene expression, we calculated p-values and FDR values by Student’s two-tailed, heteroscedastic t-test, adjusting for false discovery using the Benjamini-Yekutieli method. We maintained COVID-19 severity groups as outlined in the original manuscript (10). Data and R statistical analysis methods generated by Shen et al. were downloaded from https://github.com/guomics-lab/CVDSBA and run in accordance with the original paper (11).

### Ingenuity Pathway Analysis

Differential gene expression values from the advanced analysis performed by the nSolver software, NanoString Technologies, Inc. were loaded into the Ingenuity Pathway Analysis (IPA) software, Qiagen Digital Insights (Figure 2). Log2 transformed and Benjamini-Yekutieli calculated FDR values were used as input for further analyses. The “Antigen Presentation Pathway” in IPA was loaded and expanded using the grow function. All upstream regulators of CIITA and PAX5 were loaded onto the pathway. Only transcripts that were represented with in the dataset were selected, all others were removed. As CD19 is a known target of PAX5 in B cell signaling and lineage maintenance, it was included in the final pathway. The “Complement System” pathway was left unmodified.

### Cell-Type Analysis

Cell-type analysis and scoring in nSolver software was performed for each of the covariate groups (Figure 3). A significant correlation between immune cell type-specific gene set expression over all samples was used to determine the validity of the cell-type score, indicating the relative abundance of a given cell type. A significant correlation between CD8A and CD8B was used to estimate the abundance of CD8+ T cells (p=0.01), and the B cell population was estimated using a correlation between CD19 and Membrane Spanning 4-Domains A1 (MS4A1) (p=0). Cell-type scores were then log2 transformed, and gaussian statistical analysis t-tests were performed with homogeneous variances.

**Figure 3.**
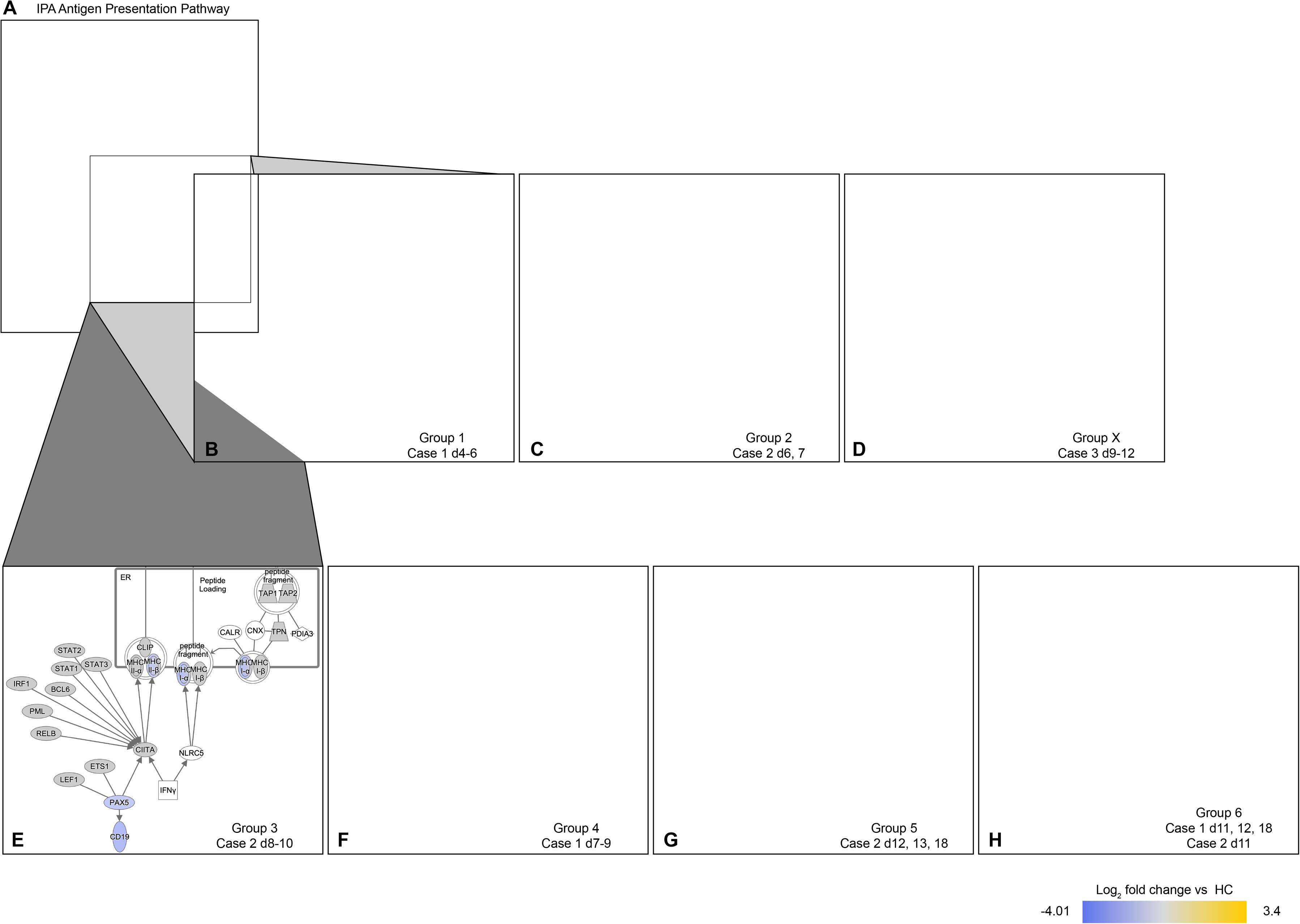
COVID-19 reduced B cell signature, specification, and lineage commitment gene abundance regardless of symptom severity. (A) Ingenuity Pathway Analysis (IPA) “Antigen Presentation Pathway” showing MHC-I and II antigen presentation pathways with the addition of genes represented in the data set which are also 1) upstream regulators of CIITA (STAT 1-3, BCL6, IRF1, PML, RELB, and PAX5), 2) upstream regulators of PAX5 (LEF1 and ETS1) and 3) a downstream target of PAX5 (CD19). (B-H) Zoomed square in (A) to show detail. Overlaid are Log2 fold-change differential expression (DE) values when compared to HC for each covariate group. All groups showed increases in STAT1 and decreases in PAX5, and CD19. Yellow and blue fill denote significantly (FDR<0.01) increased or decreased fold-regulation over HC, respectively. (B) In addition to DE in all groups, group 1 (case 1 d4-6 (severe)) showed increases in MHC-I signaling components, HLA-A, TAP1/2. Despite increased upstream regulators (STAT 1-3, BCL6, IRF1, PML), CIITA was not increased. MHC-II receptors (HLA-DMA/B/OB/PB1/RA/RB1/RB3), CLIP (CD74), LET1, and ETS1showed decreased DE. (C) Group 2 (case 2 d6, 7 (mild)) showed increased HLA-C, STAT1/2, and PML and decreases in HLA-DOB/RB. (D) Group X (Case 3 (d9-12 mild-moderate)) displayed decreased HLA-DRA abundance. (E) Group 3 showed decreased HLA-C, and HLA-DOB/RB1. (F) Group 4 showed decreased CLIP (CD74), LEF1, HLA-C, and HLA-DMB/OB/PB1/RA/RB1/DRB3. (G) The latest clinical samples for case 2 (mild) d12, 13, and 18 (group 5) showed decreased HLA-C and HLA-DOB/RA/RB1. (H) The latest clinical samples for case 1 (severe) (group 6) showed decreased RELB, LEF1, HLA-C and HLA-DMB/OB/PA1/DPB1/RA/RB1. Greyed molecules are in the dataset but do not meet the cutoff of more than 2-fold regulated with an FDR<0.01.

**Figure 4.**
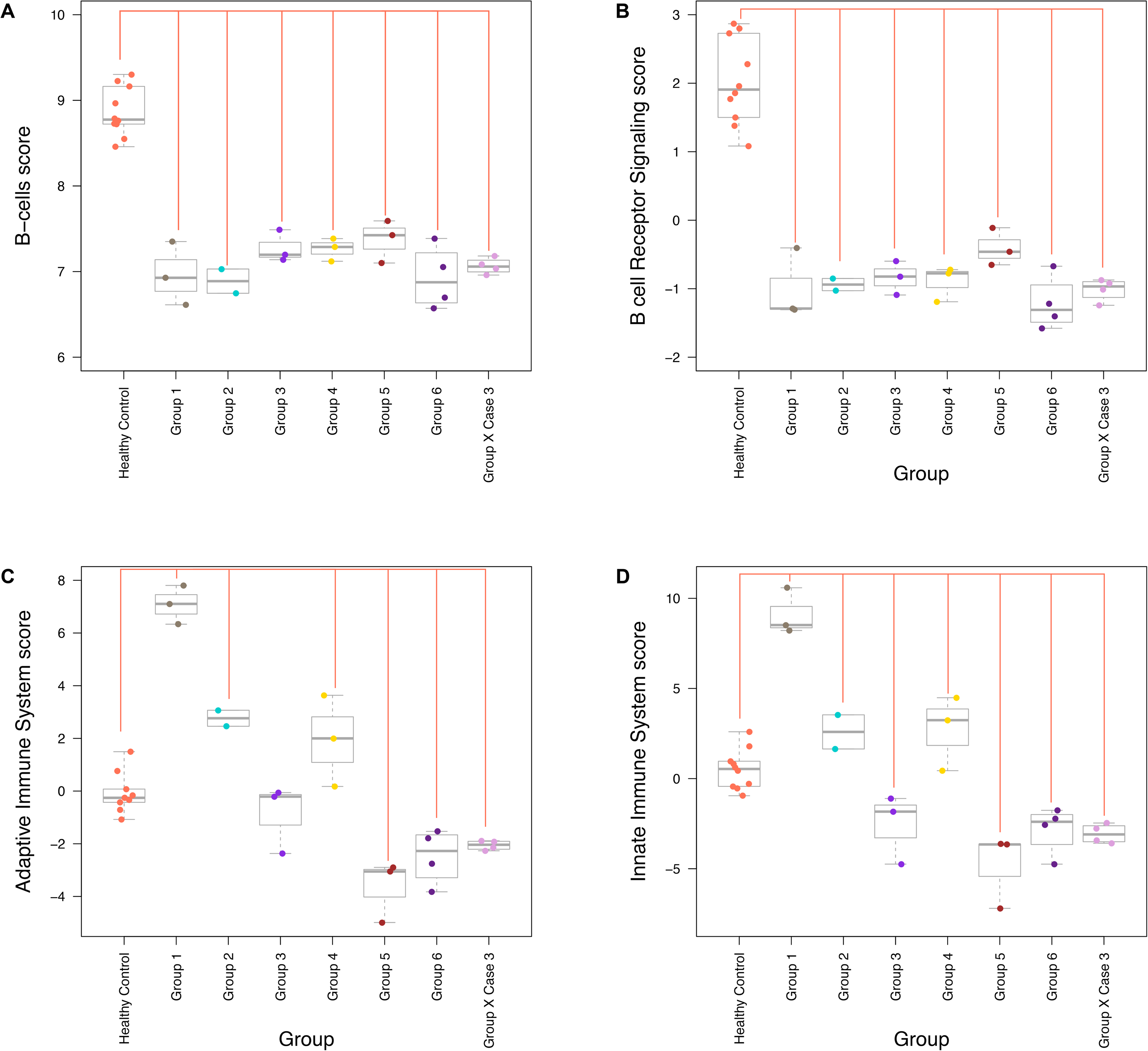
COVID-19 negatively affected estimated B cell abundance, and globally depressed adaptive and innate immune-related transcripts as SARS-CoV-2 infection resolved. (A-E) Boxplot graphs of means and variance of cell scores (A and B) or pathway scores (C-E) plotted against each group. (A) Estimating the relative abundance, CD8+ T cell scores showed a significant decrease in groups 1, 2, 4, and 6 when compared to HC. Additionally, group 2 showed a recovery of CD8+ T cells because group 3 was significantly increased. Group 6, mostly samples from the last phase of recovery of case 1 (severe), showed that CD8+ T cells did not recover in case 1. (B) B cell scores imply a significant depletion in the estimated abundance of this cell in all groups when compared to HC. There were no significant differences between group samples. (C) B cell receptor signaling aligns with estimated B cell numbers, which showed as significantly more negative than HC. There were no significant differences between group samples. (D) Although adaptive immune-related transcripts increased in groups 1, 2, and 4, this cohort of genes was significantly depressed later in the clinical course of COVID-19 groups 5, 6, and X. (E) Innate immune-related transcript scores were significantly increased in groups 1, 2, and 4 but were significantly decreased compared to HC in groups 3, 5, 6, and X, representing the later phases of COVID-19 recovery.

## Results and Discussion

### COVID-19 increased SERPING1 – a transcript central to coagulation and complement regulation

A visual inspection of the hierarchical analysis in Ong et al., where relative gene expression values were clustered by function for 10 healthy controls and 22 COVID-19 samples collected from 3 patients, showed roughly three distinct phases of gene expression in case 1, two phases in case 2, and relatively stable gene expression throughout the more limited temporal samples of case 3 (8).

To justify our statistical analysis, we took an unbiased approach to assign the clinical samples into groups and performed an unsupervised hierarchical clustering analysis (HCA) of all 32 clinical samples by similarities in global gene expression profiles (Figure 1A). Visually, the left 15 columns showed increased global immune transcript expression (higher proportion of yellow) compared to the other 17 columns (higher proportion of blue).

All healthy control (HC) samples clustered together two branch points down, and, surprisingly, group 1 (case 1 days (d)4-6) co-clustered with the HC group off of the first branch point away from all of the other case samples, implying that despite all other factors (relation, time of illness, etc.), the unweighted global immune profile during these days of infection of case 1 was more closely related to the HC group than the other COVID-19 samples. Group 1 displayed the most severe clinical symptoms of COVID-19 during this phase, with SO2 nadir on day 5. As expected, group 1 displayed a global increase in immune response-related transcripts, but, unexpectedly, this was the only cluster that had higher unweighted global gene expression levels of immune-response related transcripts than HC samples.

The rest of the clinical samples clustered together on a separate tree, with group 2 (case 2 d6 and d7) clustering the closest to the tree with group 1 and HC. The clinical course of case 2 was characterized by mild symptoms without the presence of a fever, so it may be expected that samples from this case would cluster closer to the HC group. Sharing a node two levels down with group 2 are group 3 (case 2 d8-10) and group X (all samples from case 3). The clustering of groups 3 and X implies that despite the genetic differences between these unrelated cases, the course of the viral infection was most similar between mild case 2 and mild-moderate case 3, many days after the onset of COVID-19 symptoms.

In a separate sub-cluster, group 4 (case 1 d7-9) shares a second-down branch point with group 5 (case 2 d12, d13, and d19) and group 6 (case 1 d11, d12, and d18 and case 2 d11). The distant separation of clusters corresponding to group 1 (case 1 d4-6) and group 4 (case 1 d7-9) may be due to either the effects of the antiviral medication or the natural course of infection, despite their temporal nature. It appears that the final phase of infection for both clinically distinct cases 1 and 2 begins on day 11, at which time both case 1 and case 2 d11 converge into one closely related cluster.

Principal component (PC) analysis ranked each sample by the strength of similarity and showed that groups that were defined based on hierarchical clustering also associated with each other over PC1 and PC2, which represented 31% and 22% of the variation between samples, respectively (Figure 1B). Based on the expression of genes captured by the first principal component (PC1 – x-axis), group 1 closely associated with HC samples and was positioned between HC and the rest of the COVID-19 samples, unlike in the hierarchal clustering analysis. When similarities were weighted along the second component that contributed the second-highest level of variance in gene expression between samples (PC2 – y-axis), the group 1 cluster showed the highest degree of separation from HC and group 5, which corresponds temporally to the latest samples collected from case 2 (d12-13, d19).

Group 1 (case 1 d4-6) represented the earliest samples collected from the most severe patient, and thus we selected for further analysis differentially expressed transcripts based on the significance of differential expression between HC and group 1 (p<0.01). The top 20 differentially increased and decreased transcripts in group 1 represented diverse genes encompassing several facets of innate, adaptive, and other related immune responses (S1 Table). The topmost differentially increased gene in group 1 was also significantly increased in groups 2 and X, and thus all three cases during their clinical courses experienced an increased abundance of serine protease inhibitor (serpin) family G, member 1 (SERPING1). SERPING1 is transcriptionally regulated by IFN-gamma (IFNG) activation of STAT-1 (12), and Cameron et al. showed that SERPING1 transcripts were increased in whole blood mRNA sampled from SARS-CoV-1 patients (9). Further, Hadjadj et al. reported a significant increase SERPING1 when comparing whole blood mRNA collected from 11 COVID-19 patients to 13 HC (10).

Differences were not reported for comparisons between severe and critical patients compared to HC samples; thus, we performed an independent analysis and observed a significant increase in SERPING1 abundance in all three COVID-19 severity groups, mild/moderate, severe, and critical, compared to HC (log2 fold-ratio COVID-19 vs healthy controls, FDR<0.01; mild/moderate: 5.30-fold; severe: 5.33-fold; critical: 4.81-fold).

SERPING1 RNA levels do not always mirror the abundance of C1 esterase inhibitor (C1-INH), the protein encoded by SERPING1 (13). Shen et al. characterized proteins in COVID-19 patient sera and identified C1-INH as 1 of 29 important distinguishing proteins and metabolites increased in sera from patients with severe compared to non-severe COVID-19 (11).

Additionally, C1-INH was significantly increased in non-severe COVID-19 patients when compared to healthy controls (11). C1-INH suppresses activation of complement, coagulation, inflammation, and fibrinolysis through inhibition of the contact system (14). This system is termed the contact system because the serine protease, factor XII (FXII) is activated by contact with anionic surfaces resulting in 1) the activation of the Kallikrein-Kinin pathway that drives bradykinin-associated inflammation, 2) cleavage of FXI, resulting in increased thrombin, fibrin formation, and fibrin clot formation, 3) activation of complement through C1r and C1s, and C4) promotion of fibrinolysis (14).

SARS-CoV-2 infection may be associated with hypercoagulopathy, and this possibility has become one of the major focuses of COVID-19 research (1-6). As C1-INH is the primary inhibitor of FXIIa, it broadly inhibits inflammation, coagulation, complement activation, and fibrinolysis (14). However, excess plasma levels of serpin-protein family members that also regulate FXIIa have been shown to inappropriately reduce fibrinolysis and are associated with increased risk of arterial thrombosis (15). Deficiency of C1-INH in humans causes hereditary angioedema, for which one treatment is human plasma-derived C1-INH (16). Thromboembolic events have been reported with C1-INH treatment (16). These data implicate that SERPING1-mediated increases in C1-INH in plasma may shift the balance between coagulation and fibrinolysis that may result in increased fibrin clots. If increased plasma C1-INH acts to inhibit fibrinolysis and contributes to hypercoagulopathy in COVID-19 patients, plasma C1-INH concentration may provide insight as a biomarker of thrombophilia in infected individuals, even in patients with mild symptoms and no fever, such as case 2 in our study.

SERPING1-encoded C1-INH is the only known natural inhibitor of the complement proteins C1r and C1s, such that they are unable to form the C1 complement activating complex (17). Complement activation is a first line of defense against viral infection (18), and deposition of complement proteins on the surface of SARS-CoV-1 has been shown to inhibit infection *in vitro* (19). In addition to C1-INH, Shen et al. also reported the involvement of complement proteins and metabolites in COVID-19 patients (11). Complement-associated transcripts significantly increased more than two-fold in our dataset, resulting in both a significantly increased complement pathway score for groups 1-4, 6, and X compared to HC and increases in components of the canonical IPA pathway “Complement System” (S2 Table, Figure 1C, Figure 2A and C).

SERPING1 is an IFN-stimulated gene (ISG) and acts in a negative-feedback loop to control contact and complement system activation to avoid damage to healthy host cells (20). CR1, CR3, and CR4 complexes inhibit complement activation, and components of these complexes were differentially increased in group 1 (S2 Table). Increased differential expression of complement-activating proteins C2 and C1QB (20) between HC and groups 1-3 and X implies complement activation. Notably, in the mild and moderate cases 2 and X, the complement inhibitor transcripts were either not regulated or decreased (S2 Table). We found a notable overlap between increased transcripts in cases 1 and 2 when we superimposed COVID-19 serum protein levels that were differentially abundant compared to healthy controls (FDR<0.01) (11) onto the IPA complement system pathway (Figure 2B and D). Increased levels of members of the membrane attack complex support that complement is activated in COVID-19 patients (11).

In SARS-CoV-2 patients, increased SERPING1-encoded C1-INH may be interpreted as an attempt by the host to inhibit inappropriate complement activation. Activated complement-associated microvascular injury by fibrin deposition and neutrophil permeation has been observed in severe COVID-19 patients that experienced respiratory failure and purpuric skin rashes (2). A recent pre-print showed that *in vitro* C1-INH attenuates SARS-CoV-2 nucleocapsid protein-mediated complement activation (21), and treatment of COVID-19 patients with complement inhibitors may be a viable therapeutic option (21-23).

### COVID-19 elicited interferon type-I signaling response regardless of symptom severity

Consistent with positive complement system scores, more than 50% of IFN type I (IFN-I)- and IFN-II-related transcripts increased, resulting in significantly positive pathway scores (S3 Table and Figure 1D and E). IFN signaling effectively controls viral infection; however, viruses have evolved defense mechanisms to evade IFN-mediated viral clearance (24). In bronchoalveolar lavage fluid from COVID-19 patients, ISGs were strongly increased, implying a robust IFN response (25). Both IFN-I and -II pathway scores were significantly increased compared to HC in groups 1 (severe) and 2 (mild) (Figure 1D and E). IFN responses between groups 1 and 2 were strikingly similar, with 13 shared transcripts more than two-fold increased. Furthermore, IFN-induced transmembrane family member 1 (IFITM-1), IFN induced protein 35 (IFI35), and C-X-C motif chemokine ligand 10 (CXCL10), all ISGs, were differentially increased in both groups 1 and 2. These data imply that although afebrile with a mild cough and sore throat, case 2 experienced a significant IFN response to SARS-CoV-2 infection. Consistently, Hadjadj et al. observed robust IFN-I and IFN-II responses in mild/moderate COVID-19 patients (10).

Although upregulated by IFN-γ, MHC class II receptors (HLA-DMB/OB/PB1/RA/RB1/RB3) and the MHC class II upstream transcriptional regulator CIITA were strongly decreased compared to HC in group 1 (early, severe), and at least one HLA gene was significantly decreased in all groups (S4 Table). The decrease in CIITA in group 1 was observed despite increases in several upstream inducers (PML, IRF1, BCL6, STAT 1-3); however, this increase was not seen in the upstream inducer PAX5 (Figure 2B). PAX5 is a signature B cell gene that is required for B cell lineage commitment, lineage restriction, and maturation (26, 27), and inhibition of PAX5 transcription reduces its target gene, CD19, and reprograms mature B cells into macrophages (28). Although not observed in Hadjadj et al. (10), both PAX5 and CD19 were significantly decreased in all of the COVID-19 sample groups, regardless of genetic background or clinical severity, and levels had not recovered by 18 or 19 DSO in cases 1 and 2 (group 5 (case 2 d12, d13, d19) and group 6 (case 1 d11, d12, d18 and case 2 d11)) (Figure 2C-H).

### COVID-19 negatively affected estimated B cell abundance, and globally depressed adaptive and innate immune-related transcripts as SARS-CoV-2 infection resolved

Consistent with the patterns of expression of PAX5 and CD19, there was a striking depletion of estimated numbers of B cells in all COVID-19 groups comprising clinical samples from all three cases, implying that B cell populations were negatively affected regardless of COVID-19 infection severity (Figure 3A). Considering that in the mild COVID-19 case 2 the estimated B cell abundance had not recovered by 19 DSO, it would have been of great clinical interest to have determined whether B cell populations had recovered by 29 DSO. Pathway scores for B cell signaling were negative compared to HC over all groups (Figure 3B). Although increases in adaptive and innate immune system-related transcripts occurred in groups 1 and 2, these gene sets were significantly decreased compared to HC in the later phases of recovery for groups 5, 6, and X, implying global suppression of innate and adaptive immune responses as SARS-CoV-2 infection resolves (Figure 3C and D).

It is tempting to speculate that mild case 2 may have had an extended viral load because of an insufficient humoral immune response, and in a comparison study, asymptomatic COVID-19 patients displayed a prolonged SARS-CoV-2 viral load and decreased levels of viral specific IgG versus those that were symptomatic (29). Extended viral shedding has been observed for SARS-CoV-1, Middle East Respiratory Syndrome (MERS), and SARS-CoV-2 (29-31). Remarkably, SARS-CoV-1 viral RNA and SARS-CoV-1-specific antibodies have been found simultaneously in plasma from patients with varying disease severities (31). Moreover, urine samples collected from such patients were able to infect susceptible kidney cells *in vitro* (31). These data call into question whether SARS-specific antibodies mount an immune response sufficient to protect against reinfection. Furthermore, there remains a question as to how long SARS-CoV-2 immunity lasts, as there has now been a confirmed case of reinfection of an individual with a strain of SARS-CoV-2 distinct from his original infection, just 4.5 months later (32).

### Concluding statements

Although these data are from a limited number of patients, we observed remarkably parallel immune responses in case 1 and his son, case 2, despite starkly different clinical courses. Case 1 was hospitalized for an extended period and required supplemental oxygen, whereas his son’s symptoms were mild, and he was without fever over 29 days of SARS-CoV-2 infection.

All three cases showed signs of global immune depression, and we speculate that a potential dramatic increase of SERPING1-encoded C1-INH may drive increased fibrin clot formation as well as global immune depression and may be viewed as a biological response to the complement hyperactivation associated with COVID-19, even in mild cases. Under other circumstances, case 2 may have dismissed his mild cough and sore throat as seasonal allergies, a cold, or air-quality issues in the absence of a fever and never realized he was positive for SARS-CoV-2. Currently there is an extreme paucity of information regarding the immune response over the course of infection in individuals that have very mild symptoms, as other related clinical studies have inclusion criteria that include fever (10, 33). Our study showed that a surprising number of diverse immune and inflammatory pathways were activated or repressed in afebrile case 2.

To our knowledge, we are the first to propose a connection between SERPING1-encoded C1-INH, complement activation, and hypercoagulation, even in patients with very mild COVID-19 symptoms. Furthermore, a hyper-response to aberrant complement activation by the extreme upregulation of SERPING1-encoded C1-INH may lead to widespread immune depression, similar to what is observed in patients with systemic inflammatory syndrome (34). Consideration must be given to the long-term effects of the dysregulation of these pathways in COVID-19 patients, especially those with potentiating comorbidities.

## Data Availability

All original data are available and uploaded to Mendeley.com.

https://data.mendeley.com/datasets/sbv5rnmp8t/draft?a=d8bedf37-2782-49b9-aea6-e8589503057b

## Acknowledgments

The authors would like to thank Rick Calvo for his input into the hierarchical clustering and principal component analyses used in this study and Erica Sercy for proofreading and editing of the manuscript.

## Supporting information

**S1 Table.**
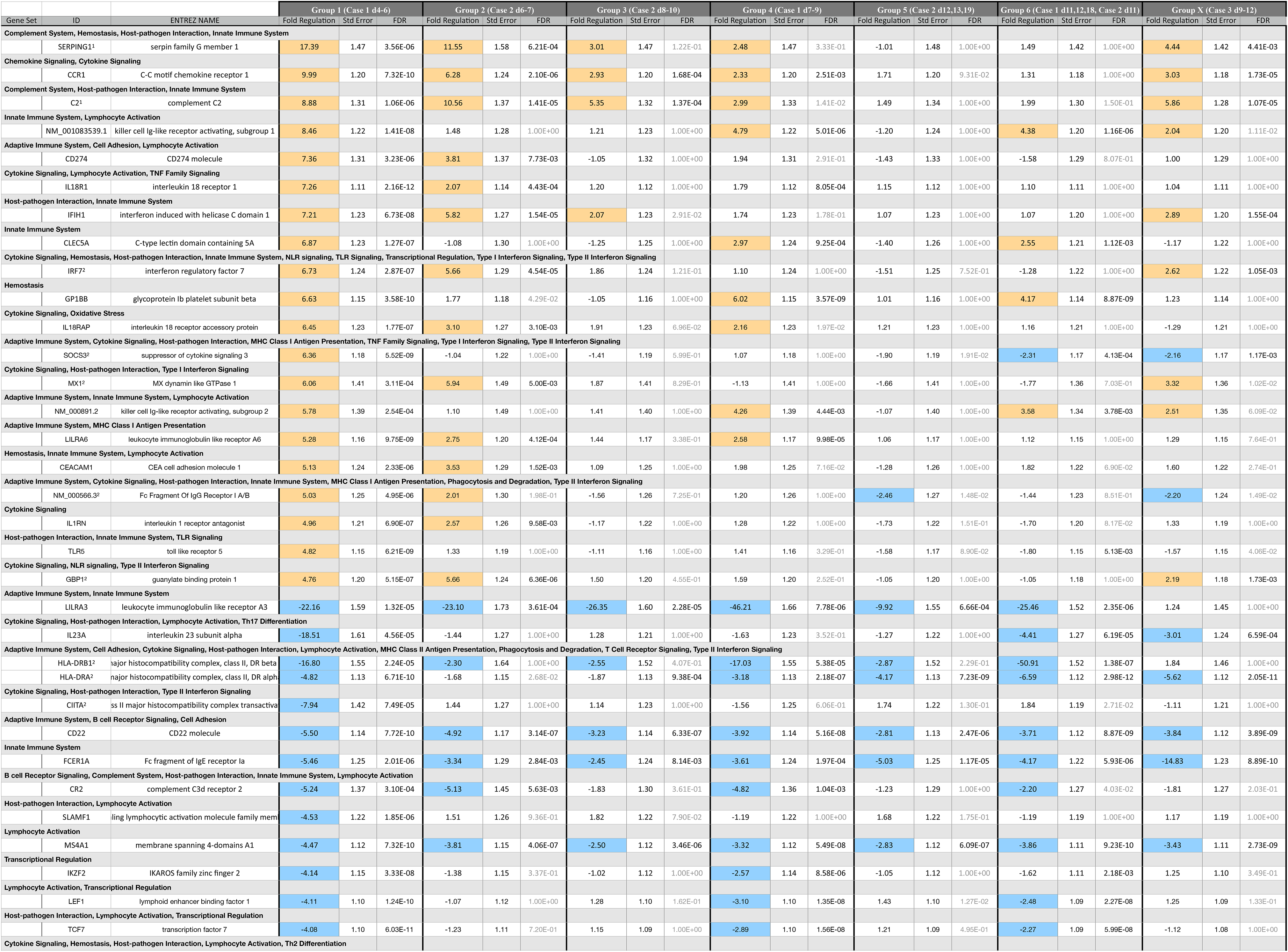
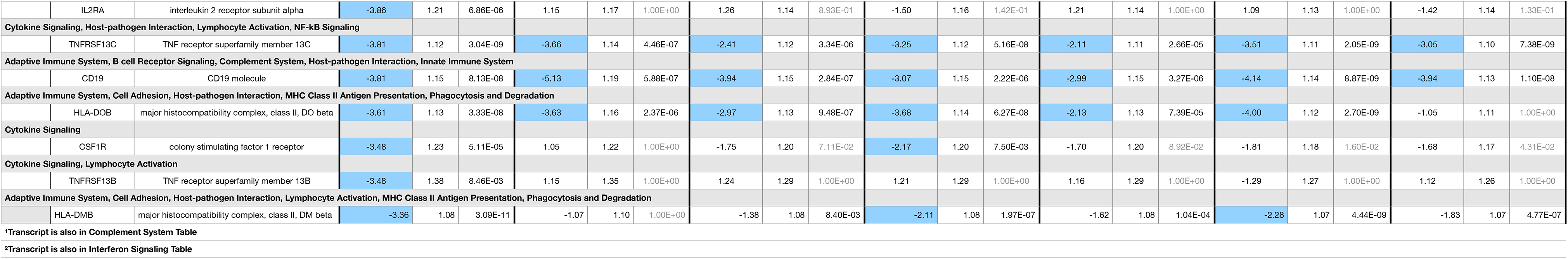
Top 20 Differentially Expressed Genes According to Group 1. Table shows the 20 most upregulated or downregulated genes in group one compared to healthy control group. Genes are categorized in “gene sets,” which are transcripts grouped together by the NanoString, Inc. software, nSolver on data in the literature that supports the involvement of a gene set in a given signaling pathway to facilitate a higher level of interpretation of the dataset as a whole. All associated gene set pathways are listed above each transcript. Selection for further analysis of differentially expressed transcripts was based on the significance of differential expression between HC and group 1 (FDR<0.01). Fold-regulation, standard error, and FDR are shown for each group. Differentially expressed transcripts that were greater than two-fold increased are highlighted orange, whereas greater than two-fold decreased transcripts are highlighted in blue. Insignificant p-values (over p=0.01) in groups 2-6 and X are denoted by grey text. lGene is also listed in the table showing the complement system gene set. 2Gene is also listed in the table showing the interferon signaling gene set.

**S2 Table.** Complement System Gene Set. Transcripts listed comprise the NanoString Technologies, Inc. “gene set” for the complement system and are categorized by all other associated gene sets. The table is sorted in descending order according to differential regulation in group 1. Fold-regulation, standard error, and FDR are shown for each group.

|  |  |  | Group 1 (Case 1 d4-6) |  |  | Group 2 (Case 2 d6-7) |  |  | Group 3 (Case 2 d8-10) |  |  | Group 4 (Case 1 d7-9) |  |  | Group 5 (Case 2 d12,13,19) |  |  | Group 6 (Case 1 d11,12,18, Case 2 d11) |  |  | Group X (Case 3 d9-12) |  |  |
| --- | --- | --- | --- | --- | --- | --- | --- | --- | --- | --- | --- | --- | --- | --- | --- | --- | --- | --- | --- | --- | --- | --- | --- |
| Gene Set | ID | ENTREZ NAME | Fold Regulation | Std Error | FDR | Fold Regulation | Std Error | FDR | Fold Regulation | Std Error | FDR | Fold Regulation | Std Error | FDR | Fold Regulation | Std Error | FDR | Fold Regulation | Std Error | FDR | Fold Regulation | Std Error | FDR |
| Hemostasis, Host-pathogen Interaction, Innate Immune System |  |  |  |  |  |  |  |  |  |  |  |  |  |  |  |  |  |  |  |  |  |  |  |
|  | SERPING1 | serpin family G member 1 | 17.39 | 1.47 | 3.56E-06 | 11.55 | 1.58 | 6.21E-04 | 3.01 | 1.47 | 1.22E-01 | 2.48 | 1.47 | 3.33E-01 | -1.01 | 1.48 | 1.00E+00 | 1.49 | 1.42 | 1.00E+00 | 4.44 | 1.42 | 4.41E-03 |
| Host-pathogen Interaction, Innate Immune System |  |  |  |  |  |  |  |  |  |  |  |  |  |  |  |  |  |  |  |  |  |  |  |
|  | C2 | complement C2 | 8.88 | 1.31 | 1.06E-06 | 10.56 | 1.37 | 1.41E-05 | 5.35 | 1.32 | 1.37E-04 | 2.99 | 1.33 | 1.41E-02 | 1.49 | 1.34 | 1.00E+00 | 1.99 | 1.30 | 1.50E-01 | 5.86 | 1.28 | 1.07E-05 |
|  | CR1 | complement C3b/C4b receptor 1 (Knops blood group) | 3.51 | 1.15 | 1.28E-07 | -1.33 | 1.17 | 9.33E-01 | -1.48 | 1.15 | 1.17E-01 | 1.73 | 1.15 | 9.80E-03 | -1.36 | 1.15 | 3.76E-01 | -1.16 | 1.13 | 1.00E+00 | 1.00 | 1.13 | 1.00E+00 |
|  | C1QB | complement C1q B chain | 3.41 | 1.33 | 2.57E-03 | 6.06 | 1.38 | 4.43E-04 | 2.89 | 1.33 | 1.96E-02 | 1.73 | 1.34 | 7.54E-01 | -1.34 | 1.38 | 1.00E+00 | 1.05 | 1.32 | 1.00E+00 | 2.62 | 1.30 | 1.55E-02 |
| Adaptive Immune System, Cell Adhesion, Cytokine Signaling, Hemostasis, Host-pathogen Interaction, Innate Immune System, Lymphocyte Activation, Lymphocyte Trafficking, Phagocytosis and Degradation, TLR Signaling |  |  |  |  |  |  |  |  |  |  |  |  |  |  |  |  |  |  |  |  |  |  |  |
|  | ITGB2 | integrin subunit beta 2 | 3.12 | 1.07 | 5.04E-12 | 1.74 | 1.08 | 2.22E-05 | 1.50 | 1.07 | 1.22E-04 | 2.10 | 1.07 | 1.56E-08 | 1.21 | 1.07 | 1.07E-01 | 1.40 | 1.06 | 1.93E-04 | 1.42 | 1.06 | 1.19E-04 |
| Cell Adhesion, Cytokine Signaling, Hemostasis, Host-pathogen Interaction, Innate Immune System, Lymphocyte Trafficking, Phagocytosis and Degradation, TLR Signaling |  |  |  |  |  |  |  |  |  |  |  |  |  |  |  |  |  |  |  |  |  |  |  |
|  | ITGAM | integrin subunit alpha M | 2.77 | 1.07 | 7.57E-11 | 1.00 | 1.09 | 1.00E+00 | -1.30 | 1.07 | 2.43E-02 | 1.66 | 1.07 | 1.43E-05 | -1.41 | 1.07 | 1.91E-03 | 1.22 | 1.07 | 5.17E-02 | -1.19 | 1.07 | 1.55E-01 |
| Cell Adhesion, Cytokine Signaling, Hemostasis, Host-pathogen Interaction, Innate Immune System |  |  |  |  |  |  |  |  |  |  |  |  |  |  |  |  |  |  |  |  |  |  |  |
|  | ITGAX | integrin subunit alpha X | 2.31 | 1.13 | 8.72E-06 | 1.11 | 1.15 | 1.00E+00 | -1.39 | 1.13 | 1.59E-01 | 1.42 | 1.13 | 1.15E-01 | -1.42 | 1.13 | 1.11E-01 | -1.18 | 1.12 | 1.00E+00 | -1.71 | 1.12 | 1.09E-03 |
| Hemostasis, Innate Immune System |  |  |  |  |  |  |  |  |  |  |  |  |  |  |  |  |  |  |  |  |  |  |  |
|  | PLAUR | plasminogen activator, urokinase receptor | 1.68 | 1.13 | 4.16E-03 | 1.10 | 1.16 | 1.00E+00 | -1.66 | 1.14 | 1.74E-02 | 1.20 | 1.14 | 1.00E+00 | -1.21 | 1.14 | 1.00E+00 | -1.44 | 1.13 | 6.22E-02 | -1.38 | 1.13 | 1.37E-01 |
| Innate Immune System, Lymphocyte Activation |  |  |  |  |  |  |  |  |  |  |  |  |  |  |  |  |  |  |  |  |  |  |  |
|  | CD59 | CD59 molecule (CD59 blood group) | 1.31 | 1.06 | 6.10E-04 | -1.19 | 1.07 | 1.61E-01 | -1.42 | 1.06 | 5.44E-05 | -1.25 | 1.06 | 7.92E-03 | -1.38 | 1.06 | 1.58E-04 | -1.22 | 1.05 | 5.05E-03 | -1.46 | 1.05 | 2.60E-06 |
| Hemostasis, Host-pathogen Interaction |  |  |  |  |  |  |  |  |  |  |  |  |  |  |  |  |  |  |  |  |  |  |  |
|  | C1QBP | complement C1q binding protein | 1.25 | 1.06 | 4.58E-03 | 1.31 | 1.07 | 6.61E-03 | 1.28 | 1.06 | 3.61E-03 | 1.14 | 1.06 | 3.14E-01 | 1.10 | 1.06 | 7.52E-01 | -1.15 | 1.05 | 7.81E-02 | 1.33 | 1.05 | 1.17E-04 |
| Adaptive Immune System, B cell Receptor Signaling, Host-pathogen Interaction, Innate Immune System |  |  |  |  |  |  |  |  |  |  |  |  |  |  |  |  |  |  |  |  |  |  |  |
|  | CD19 | CD19 molecule | -3.81 | 1.15 | 8.13E-08 | -5.13 | 1.19 | 5.88E-07 | -3.94 | 1.15 | 2.84E-07 | -3.07 | 1.15 | 2.22E-06 | -2.99 | 1.15 | 3.27E-06 | -4.14 | 1.14 | 8.87E-09 | -3.94 | 1.13 | 1.10E-08 |
| B cell Receptor signaling, Host-pathogen Interaction, Innate Immune System, Lymphocyte Activation |  |  |  |  |  |  |  |  |  |  |  |  |  |  |  |  |  |  |  |  |  |  |  |
|  | CR2 | complement C3d receptor 2 | -5.24 | 1.37 | 3.10E-04 | -5.13 | 1.45 | 5.63E-03 | -1.83 | 1.30 | 3.61E-01 | -4.82 | 1.36 | 1.04E-03 | -1.23 | 1.29 | 1.00E+00 | -2.20 | 1.27 | 4.03E-02 | -1.81 | 1.27 | 2.03E-01 |

**S3 Table.**
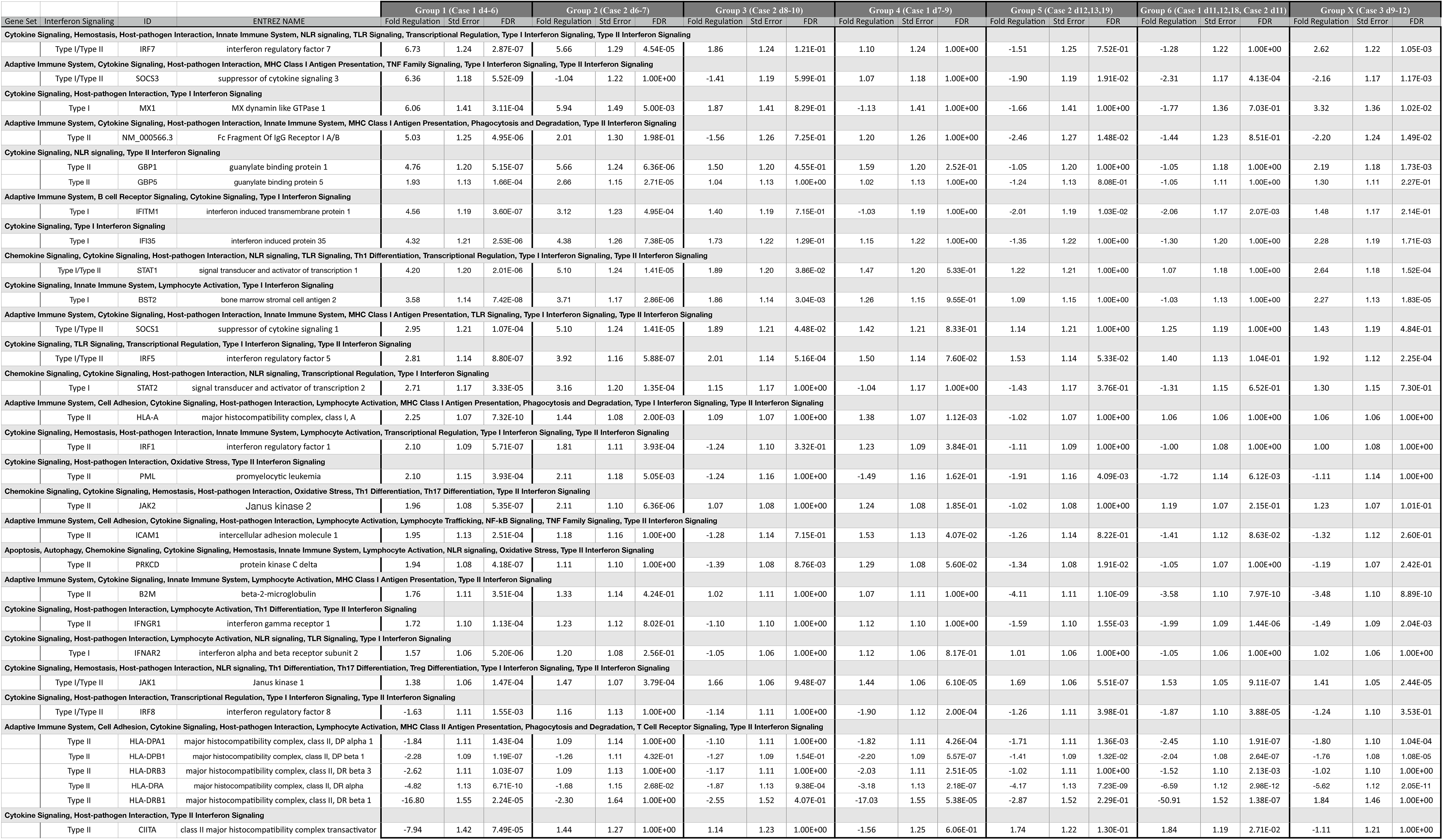
Interferon Type-I and -II Signaling Gene Sets. Genes listed comprise the NanoString Technologies, Inc. “gene set” for Interferon Type I and II signaling and are categorized by all other associated gene sets. The “type” of interferon signaling according to NanoString Technologies, Inc. is listed. The table is sorted in descending order according to differential regulation in group 1. Fold-regulation, standard error, and FDR are shown sorted by group.

**S4 Table.** MHC II signaling and CIITA regulating genes referenced in Figure 2. All HLA transcripts from the NanoString Technologies, Inc. human immunology V2 array are listed along with genes identified by ingenuity pathway analysis as 1) upstream regulators of CIITA (STAT 1-3, BCL6, IRF1, PML, RELB, and PAX5), 2) upstream regulators of PAX5 (LEF1 and ETS1) and 3) a downstream target of PAX5 (CD19), which were also represented in the dataset.

|  |  |  | Group 1 (Case 1 d4-6) |  |  | Group 2 (Case 2 d6-7) |  |  | Group 3 (Case 2 d8-10) |  |  | Group 4 (Case 1 d7-9) |  |  | Group 5 (Case 2 d12,13,19) |  |  | Group 6 (Case 1 d11,12,18, Case 2 d11) |  |  | Group X (Case 3 d9-12) |  |  |
| --- | --- | --- | --- | --- | --- | --- | --- | --- | --- | --- | --- | --- | --- | --- | --- | --- | --- | --- | --- | --- | --- | --- | --- |
| Gene Set | ID | ENTREZ NAME | Fold Regulation | Std Error | FDR | Fold Regulation | Std Error | FDR | Fold Regulation | Std Error | FDR | Fold Regulation | Std Error | FDR | Fold Regulation | Std Error | FDR | Fold Regulation | Std Error | FDR | Fold Regulation | Std Error | FDR |
| Cytokine Signaling, Lymphocyte Activation |  |  |  |  |  |  |  |  |  |  |  |  |  |  |  |  |  |  |  |  |  |  |  |
|  | BCL6 | BCL6 transcription repressor | 1.78 | 0.20 | 2.45E-07 | -0.07 | 0.24 | 1.00E+00 | -0.82 | 0.20 | 1.06E-02 | 0.89 | 0.20 | 4.37E-03 | -0.73 | 0.20 | 2.41E-02 | -0.07 | 0.18 | 1.00E+00 | -0.59 | 0.18 | 4.56E-02 |
| Adaptive Immune System, B cell Receptor Signaling, Complement System, Host-pathogen Interaction, Innate Immune System |  |  |  |  |  |  |  |  |  |  |  |  |  |  |  |  |  |  |  |  |  |  |  |
|  | CD19 | CD19 molecule | -1.93 | 0.20 | 8.13E-08 | -2.36 | 0.25 | 5.88E-07 | -1.98 | 0.20 | 2.84E-07 | -1.62 | 0.20 | 2.22E-06 | -1.58 | 0.20 | 3.27E-06 | -2.05 | 0.18 | 8.87E-09 | -1.98 | 0.18 | 1.10E-08 |
| Adaptive Immune System, Hemostasis, Host-pathogen Interaction, Lymphocyte Activation, MHC Class II Antigen Presentation |  |  |  |  |  |  |  |  |  |  |  |  |  |  |  |  |  |  |  |  |  |  |  |
|  | CD74 | CD74 molecule | -1.24 | 0.11 | 3.00E-09 | -0.24 | 0.12 | 7.20E-01 | -0.55 | 0.11 | 8.25E-04 | -1.01 | 0.11 | 1.97E-07 | -0.48 | 0.11 | 2.72E-03 | -0.89 | 0.09 | 1.37E-07 | -0.86 | 0.09 | 2.17E-07 |
| Cytokine Signaling, Host-pathogen Interaction, Type II Interferon Signaling |  |  |  |  |  |  |  |  |  |  |  |  |  |  |  |  |  |  |  |  |  |  |  |
|  | CIITA | class II major histocompatibility complex transactivator | -2.99 | 0.51 | 7.49E-05 | 0.52 | 0.34 | 1.00E+00 | 0.19 | 0.30 | 1.00E+00 | -0.65 | 0.32 | 6.06E-01 | 0.80 | 0.29 | 1.30E-01 | 0.88 | 0.26 | 2.71E-02 | -0.15 | 0.28 | 1.00E+00 |
| Host-pathogen Interaction, Oxidative Stress, Transcriptional Regulation |  |  |  |  |  |  |  |  |  |  |  |  |  |  |  |  |  |  |  |  |  |  |  |
|  | ETS1 | ETS proto-oncogene 1, transcription factor | -1.02 | 0.10 | 3.51E-08 | -0.28 | 0.12 | 3.16E-01 | 0.04 | 0.10 | 1.00E+00 | -0.93 | 0.10 | 3.36E-07 | 0.05 | 0.10 | 1.00E+00 | -0.75 | 0.09 | 8.98E-07 | -0.28 | 0.09 | 6.72E-02 |
| Adaptive Immune System, Cell Adhesion, Cytokine Signaling, Host-pathogen Interaction, Lymphocyte Activation, MHC Class I Antigen Presentation, Phagocytosis and Degradation, Type I Interferon Signaling, Type II Interferon Signaling |  |  |  |  |  |  |  |  |  |  |  |  |  |  |  |  |  |  |  |  |  |  |  |
|  | HLA-A | major histocompatibility complex, class I, A | 1.17 | 0.09 | 7.32E-10 | 0.53 | 0.11 | 2.00E-03 | 0.13 | 0.09 | 1.00E+00 | 0.47 | 0.09 | 1.12E-03 | -0.03 | 0.09 | 1.00E+00 | 0.09 | 0.08 | 1.00E+00 | 0.08 | 0.08 | 1.00E+00 |
| Adaptive Immune System, Cell Adhesion, Cytokine Signaling, Host-pathogen Interaction, Innate Immune System, Lymphocyte Activation, MHC Class I Antigen Presentation, Phagocytosis and Degradation, Type I Interferon Signaling, Type II Interferon Signaling |  |  |  |  |  |  |  |  |  |  |  |  |  |  |  |  |  |  |  |  |  |  |  |
|  | HLA-B | major histocompatibility complex, class I, B | 0.27 | 0.10 | 1.04E-01 | -0.13 | 0.12 | 1.00E+00 | -0.50 | 0.10 | 1.19E-03 | -0.40 | 0.10 | 1.03E-02 | -0.52 | 0.10 | 7.56E-04 | -0.75 | 0.09 | 6.29E-07 | -0.46 | 0.09 | 6.09E-04 |
|  | HLA-C | major histocompatibility complex, class I, C | -0.39 | 0.15 | 1.69E-01 | -1.20 | 0.18 | 5.13E-05 | -1.76 | 0.15 | 1.74E-08 | -1.15 | 0.15 | 8.06E-06 | -1.32 | 0.15 | 1.22E-06 | -1.26 | 0.14 | 1.73E-07 | -0.48 | 0.14 | 2.64E-02 |
| Cell Adhesion, Host-pathogen Interaction, MHC Class II Antigen Presentation, Phagocytosis and Degradation |  |  |  |  |  |  |  |  |  |  |  |  |  |  |  |  |  |  |  |  |  |  |  |
|  | HLA-DMA | major histocompatibility complex, class II, DM alpha | -1.18 | 0.12 | 3.22E-08 | 0.05 | 0.13 | 1.00E+00 | -0.22 | 0.11 | 5.89E-01 | -0.63 | 0.11 | 2.95E-04 | 0.13 | 0.11 | 1.00E+00 | -0.39 | 0.10 | 8.31E-03 | -0.32 | 0.10 | 4.65E-02 |
| Adaptive Immune System, Cell Adhesion, Host-pathogen Interaction, Lymphocyte Activation, MHC Class II Antigen Presentation, Phagocytosis and Degradation |  |  |  |  |  |  |  |  |  |  |  |  |  |  |  |  |  |  |  |  |  |  |  |
|  | HLA-DMB | major histocompatibility complex, class II, DM beta | -1.75 | 0.11 | 3.09E-11 | -0.10 | 0.13 | 1.00E+00 | -0.46 | 0.11 | 8.40E-03 | -1.08 | 0.11 | 1.97E-07 | -0.69 | 0.11 | 1.04E-04 | -1.19 | 0.10 | 4.44E-09 | -0.87 | 0.10 | 4.77E-07 |
| Adaptive Immune System, Cell Adhesion, Host-pathogen Interaction, MHC Class II Antigen Presentation, Phagocytosis and Degradation |  |  |  |  |  |  |  |  |  |  |  |  |  |  |  |  |  |  |  |  |  |  |  |
|  | HLA-DOB | major histocompatibility complex, class II, DO beta | -1.85 | 0.18 | 3.33E-08 | -1.86 | 0.22 | 2.37E-06 | -1.57 | 0.18 | 9.48E-07 | -1.88 | 0.18 | 6.27E-08 | -1.09 | 0.17 | 7.39E-05 | -2.00 | 0.17 | 2.70E-09 | -0.07 | 0.15 | 1.00E+00 |
| Adaptive Immune System, Cell Adhesion, Cytokine Signaling, Host-pathogen Interaction, Lymphocyte Activation, MHC Class II Antigen Presentation, Phagocytosis and Degradation, T Cell Receptor Signaling, Type II Interferon Signaling |  |  |  |  |  |  |  |  |  |  |  |  |  |  |  |  |  |  |  |  |  |  |  |
|  | HLA-DPA1 | major histocompatibility complex, class II, DP alpha 1 | -0.88 | 0.16 | 1.43E-04 | 0.13 | 0.18 | 1.00E+00 | -0.13 | 0.16 | 1.00E+00 | -0.86 | 0.16 | 4.26E-04 | -0.78 | 0.16 | 1.36E-03 | -1.29 | 0.14 | 1.91E-07 | -0.84 | 0.14 | 1.04E-04 |
|  | HLA-DPB1 | major histocompatibility complex, class II, DP beta 1 | -1.19 | 0.13 | 1.19E-07 | -0.34 | 0.15 | 4.32E-01 | -0.35 | 0.13 | 1.54E-01 | -1.14 | 0.13 | 5.57E-07 | -0.49 | 0.13 | 1.32E-02 | -1.03 | 0.12 | 2.64E-07 | -0.81 | 0.12 | 1.08E-05 |
|  | HLA-DRA | major histocompatibility complex, class II, DR alpha | -2.27 | 0.18 | 6.71E-10 | -0.75 | 0.21 | 2.68E-02 | -0.90 | 0.18 | 9.38E-04 | -1.67 | 0.18 | 2.18E-07 | -2.06 | 0.18 | 7.23E-09 | -2.72 | 0.16 | 2.98E-12 | -2.49 | 0.16 | 2.05E-11 |
|  | HLA-DRB1 | major histocompatibility complex, class II, DR beta 1 | -4.07 | 0.63 | 2.24E-05 | -1.20 | 0.72 | 1.00E+00 | -1.35 | 0.61 | 4.07E-01 | -4.09 | 0.63 | 5.38E-05 | -1.52 | 0.61 | 2.29E-01 | -5.67 | 0.61 | 1.38E-07 | 0.88 | 0.54 | 1.00E+00 |
|  | HLA-DRB3 | major histocompatibility complex, class II, DR beta 3 | -1.39 | 0.15 | 1.03E-07 | 0.13 | 0.17 | 1.00E+00 | -0.23 | 0.15 | 1.00E+00 | -1.02 | 0.15 | 2.51E-05 | -0.03 | 0.15 | 1.00E+00 | -0.61 | 0.13 | 2.13E-03 | -0.03 | 0.13 | 1.00E+00 |
| Cytokine Signaling, Hemostasis, Host-pathogen Interaction, Innate Immune System, Lymphocyte Activation, Transcriptional Regulation, Type I Interferon Signaling, Type II Interferon Signaling |  |  |  |  |  |  |  |  |  |  |  |  |  |  |  |  |  |  |  |  |  |  |  |
|  | IRF1 | interferon regulatory factor 1 | 1.07 | 0.13 | 5.71E-07 | 0.86 | 0.15 | 3.93E-04 | -0.31 | 0.13 | 3.32E-01 | 0.29 | 0.13 | 3.84E-01 | -0.15 | 0.13 | 1.00E+00 | -0.01 | 0.12 | 1.00E+00 | 0.00 | 0.12 | 1.00E+00 |
| Lymphocyte Activation, Transcriptional Regulation |  |  |  |  |  |  |  |  |  |  |  |  |  |  |  |  |  |  |  |  |  |  |  |
|  | LEF1 | lymphoid enhancer binding factor 1 | -2.04 | 0.14 | 1.24E-10 | -0.10 | 0.16 | 1.00E+00 | 0.36 | 0.13 | 1.62E-01 | -1.63 | 0.14 | 1.35E-08 | 0.52 | 0.13 | 1.27E-02 | -1.31 | 0.12 | 2.27E-08 | 0.33 | 0.12 | 1.33E-01 |
| Transcriptional Regulation |  |  |  |  |  |  |  |  |  |  |  |  |  |  |  |  |  |  |  |  |  |  |  |
|  | PAX5 | paired box 5 | -1.75 | 0.20 | 3.21E-07 | -2.12 | 0.25 | 2.84E-06 | -1.52 | 0.20 | 5.49E-06 | -1.35 | 0.19 | 2.05E-05 | -1.12 | 0.19 | 1.58E-04 | -1.64 | 0.18 | 1.50E-07 | -1.76 | 0.18 | 6.18E-08 |
| Cytokine Signaling, Host-pathogen Interaction, Oxidative Stress, Type II Interferon Signaling |  |  |  |  |  |  |  |  |  |  |  |  |  |  |  |  |  |  |  |  |  |  |  |
|  | PML | promyelocytic leukemia | 1.07 | 0.21 | 3.93E-04 | 1.08 | 0.24 | 5.05E-03 | -0.31 | 0.21 | 1.00E+00 | -0.58 | 0.21 | 1.62E-01 | -0.94 | 0.21 | 4.09E-03 | -0.78 | 0.19 | 6.12E-03 | -0.15 | 0.19 | 1.00E+00 |
| Cytokine Signaling, Host-pathogen Interaction, Innate Immune System, Lymphocyte Activation, NF-kB Signaling, Transcriptional Regulation |  |  |  |  |  |  |  |  |  |  |  |  |  |  |  |  |  |  |  |  |  |  |  |
|  | RELB | RELB proto-oncogene, NF-kB subunit | 0.47 | 0.16 | 7.05E-02 | -0.20 | 0.20 | 1.00E+00 | -0.85 | 0.18 | 1.83E-03 | 0.14 | 0.16 | 1.00E+00 | -0.82 | 0.18 | 2.18E-03 | -1.20 | 0.16 | 4.78E-06 | -0.64 | 0.15 | 5.53E-03 |
| Chemokine Signaling, Cytokine Signaling, Host-pathogen Interaction, NLR signaling, TLR Signaling, Th1 Differentiation, Transcriptional Regulation, Type I Interferon Signaling, Type II Interferon Signaling |  |  |  |  |  |  |  |  |  |  |  |  |  |  |  |  |  |  |  |  |  |  |  |
|  | STAT1 | signal transducer and activator of transcription 1 | 2.07 | 0.27 | 2.01E-06 | 2.35 | 0.32 | 1.41E-05 | 0.92 | 0.27 | 3.86E-02 | 0.56 | 0.27 | 5.33E-01 | 0.29 | 0.27 | 1.00E+00 | 0.10 | 0.24 | 1.00E+00 | 1.40 | 0.24 | 1.52E-04 |
| Chemokine Signaling, Cytokine Signaling, Host-pathogen Interaction, NLR signaling, Transcriptional Regulation, Type I Interferon Signaling |  |  |  |  |  |  |  |  |  |  |  |  |  |  |  |  |  |  |  |  |  |  |  |
|  | STAT2 | signal transducer and activator of transcription 2 | 1.44 | 0.23 | 3.33E-05 | 1.66 | 0.27 | 1.35E-04 | 0.20 | 0.23 | 1.00E+00 | -0.06 | 0.23 | 1.00E+00 | -0.52 | 0.23 | 3.76E-01 | -0.39 | 0.21 | 6.52E-01 | 0.38 | 0.21 | 7.30E-01 |
| Chemokine Signaling, Cytokine Signaling, Host-pathogen Interaction, Th17 Differentiation, Transcriptional Regulation, Treg Differentiation |  |  |  |  |  |  |  |  |  |  |  |  |  |  |  |  |  |  |  |  |  |  |  |
|  | STAT3 | signal transducer and activator of transcription 3 | 1.21 | 0.10 | 1.03E-09 | 0.52 | 0.12 | 4.01E-03 | -0.06 | 0.10 | 1.00E+00 | 0.44 | 0.10 | 3.31E-03 | 0.02 | 0.10 | 1.00E+00 | 0.11 | 0.09 | 1.00E+00 | 0.11 | 0.09 | 1.00E+00 |
| Adaptive Immune System, Host-pathogen Interaction, MHC Class I Antigen Presentation, Phagocytosis and Degradation |  |  |  |  |  |  |  |  |  |  |  |  |  |  |  |  |  |  |  |  |  |  |  |
|  | TAP1 | transporter 1, ATP binding cassette subfamily B member | 1.16 | 0.14 | 5.44E-07 | 0.84 | 0.16 | 1.12E-03 | -0.16 | 0.14 | 1.00E+00 | 0.05 | 0.14 | 1.00E+00 | -0.76 | 0.15 | 8.37E-04 | -0.67 | 0.13 | 6.57E-04 | -0.17 | 0.13 | 1.00E+00 |
|  | TAP2 | transporter 2, ATP binding cassette subfamily B member | 1.06 | 0.13 | 9.40E-07 | 0.61 | 0.16 | 1.52E-02 | -0.24 | 0.14 | 8.60E-01 | -0.17 | 0.14 | 1.00E+00 | -0.57 | 0.14 | 6.57E-03 | -0.33 | 0.12 | 1.41E-01 | -0.69 | 0.12 | 2.19E-04 |
| Adaptive Immune System, MHC Class I Antigen Presentation |  |  |  |  |  |  |  |  |  |  |  |  |  |  |  |  |  |  |  |  |  |  |  |
|  | TAPBP (TPN) | TAP binding protein | 0.34 | 0.08 | 2.47E-03 | 0.03 | 0.09 | 1.00E+00 | -0.28 | 0.08 | 2.97E-02 | -0.19 | 0.08 | 2.69E-01 | -0.50 | 0.08 | 7.39E-05 | -0.55 | 0.07 | 2.10E-06 | -0.61 | 0.07 | 5.42E-07 |

**S5 Table.**
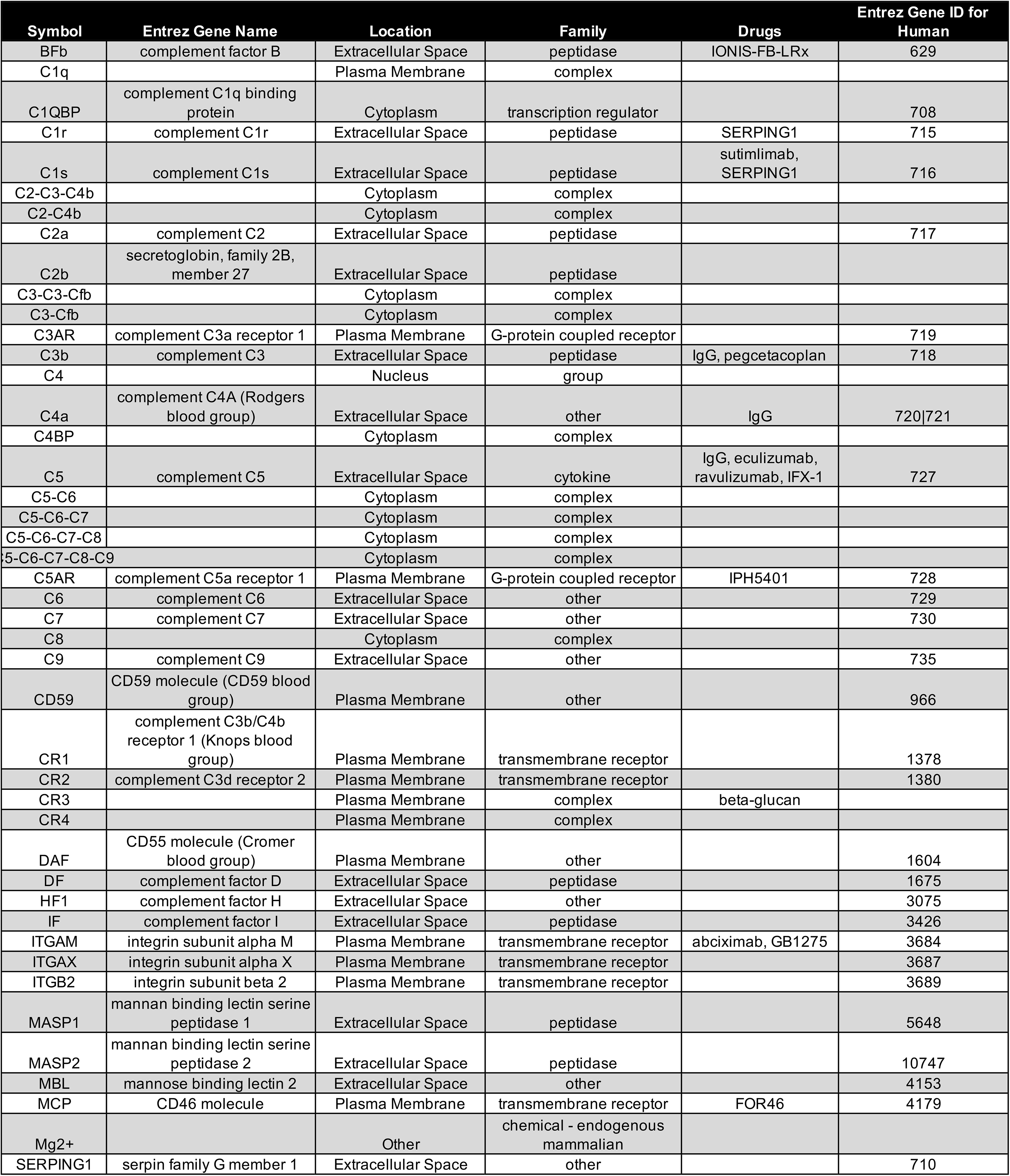
Key to IPA “Complement Scomplement ystem” genes. Columns reference the symbol used in Figure 2, the Entrez gene name, the subcellular location according to IPA, the designated molecular family according to IPA, drugs known to target that particular gene/protein, and the numerical Entrez gene ID for human.

## Notes

### Competing Interest Statement

The authors have declared no competing interest.

### Funding Statement

No external funding was received.

### Author Declarations

We have confirmed with our HCA-Healthone IRB that this study does not constitute research involving human subjects under the federal Common Rule, 45 CFR Part 46. The datasets used and/or analyzed during the current study are publicly available. Elsevier Inc. remains the full and exclusive copyright owner and is not responsible for any claims arising from works based on the original data, text, tables, or figures.

